# Stakeholder Perspectives on Adolescent Sexual and Reproductive Health Service Utilisation in Uganda’s Busoga Region: A Qualitative Exploratory Study

**DOI:** 10.1101/2025.09.22.25336327

**Authors:** Noah Nyende, Justine Nanteza, Frank Kinyingi, Miph Musoke

**Affiliations:** Nkumba University, Entebbe, Uganda; Makerere University, Kampala, Uganda

**Keywords:** Adolescent, Reproductive Health, Uganda, Qualitative Research, Health Services Accessibility

## Abstract

**Objective:** To explore adolescents’ attitudes, perceptions, and preferences regarding sexual and reproductive health (SRH) services in Busoga, Uganda, and to examine community leaders’ and health providers’ perspectives to inform youth-friendly interventions.

**Design:** Exploratory qualitative study under a constructivist paradigm, using key informant interviews (KIIs) and focus group discussions (FGDs). Analysis was inductive, following Braun and Clarke’s six-step thematic approach, and guided by the Health Belief Model and Andersen’s Behavioural Model.

**Setting:** Rural and urban sites in Iganga and Bugweri districts, Uganda.

**Participants:** Six KIIs with community leaders and six with health providers; four FGDs with adolescents (n=39).

**Results:** Five themes emerged: (1) positive attitudes toward SRH but persistent misconceptions; (2) structural and interpersonal barriers including distance, cost, and judgemental providers; (3) preference for free, peer-led, school/community-based services with gender-matched staff; (4) sociocultural constraints such as early marriage and religious prohibitions; and (5) system gaps including commodity shortages and limited staff training.

**Conclusion:** Adolescents are motivated to protect their SRH but face multilevel barriers. Culturally sensitive, adolescent-friendly strategies, peer engagement, and reliable supply chains are urgently needed.

**Key questions:** *What is already known on this topic:*

- Adolescents in sub-Saharan Africa face stigma, cost, and distance barriers to SRH services.

*What this study adds:*

- Adolescents in Busoga prefer free, peer-led, school/community-based services and describe persistent myths about contraception.

*How this study might affect research, practice, or policy:*

- Multi-stakeholder partnerships, including faith and cultural leaders, are essential for adolescent-friendly SRH policy and programming.

## Introduction

Adolescence (10–19 years) is a critical period for establishing sexual and reproductive health (SRH) behaviours that influence lifelong wellbeing and fertility outcomes. ^1 2^ Globally, adolescents, especially in low- and middle-income countries (LMICs)continue to face major SRH challenges, including high rates of unintended pregnancy and sexually transmitted infections (STIs). In sub-Saharan Africa (SSA), structural barriers such as stigma, misconceptions, and limited access contribute to low utilisation of SRH care, and persistently high rates of teenage pregnancy and HIV.^3 4^ Kenyan and Ethiopian studies, for example, highlight transport costs, judgmental providers, and restrictive cultural norms as key barriers. ^5–8^

Uganda’s adolescent pregnancy rate is 24%, HIV prevalence among 15–19-year-olds is 2.1%, and modern contraceptive use is only 15%. ^9^ The Busoga Region is particularly affected because of poverty, early marriage norms, and limited services. ^10 11^ Despite national adolescent health guidelines,^12^ utilisation remains low due to stigma, lack of privacy, and misinformation, ^13–15^ yet little is known about how multiple stakeholders, including adolescents, community leaders, and health providers, perceive these barriers and potential solutions. This gap limits the design of locally responsive, youth-friendly interventions.

This study sought to address three questions:

1. What are adolescents’ attitudes, perceptions, and preferences regarding SRH services in Busoga?
2. How do community leaders and health providers view adolescent SRH service needs and use?
3. What multilevel barriers and facilitators can inform youth-friendly interventions?

The study was guided by the Health Belief Model ^16^ and Andersen’s Behavioural Model, ^17^ to understand how individual perceptions, enabling factors, and sociocultural contexts shape SRH service utilisation.

## Methods

### Design and setting

Adopted an exploratory qualitative design informed by an interpretivist paradigm to capture diverse stakeholder meanings and experiences. The study was conducted in November 2024 at six rural (Namirali, Kiwerere, Bulubandi, Naitandu B, Bunalwenyi C, Makandwa) and six urban (Kasokoso Central II/III, Nakavule Main, Busesa, Butende, Kagamba) sites in Iganga and Bugweri districts. The two districts were selected because they reflect contrasting health system contexts: Bugweri has limited resources and fragile health infrastructure, whereas Iganga is better resourced with stronger systems and staffing, ensuring representation of diverse conditions influencing adolescent SRH service access and utilisation in the region.

### Participants and sampling

Two participant groups were included: key community informants and adolescents.

- **Key informant interviews (KIIs):** Twelve participants were purposively selected, comprising six community leaders (three cultural and three religious) and six health providers drawn from health centres II to IV and the district hospital.
- **Focus group discussions (FGDs):** Four groups of adolescents were recruited through convenience sampling. Two groups were girls aged 13 to 17 years, and two groups were boys aged 17 to 19 years, with one rural and one urban group for each gender, giving a total of 39 participants.

Sample size was determined a priori based on established guidelines, ^18^ with data saturation confirmed after 12 KIIs and four FGDs. Inclusion criteria were adolescents aged 13–19 years residing in the study areas and leaders/providers identified by their communities or workplaces as knowledgeable about adolescent SRH services.

### Data collection

Semi-structured guides were pilot tested among similar but non-study localities. Refined tools were used to explore attitudes, perceptions, preferences, and barriers. Sessions (60–90 min) were conducted in Lusoga or English. Audio recordings were transcribed verbatim, checked against recordings for accuracy, and stored on encrypted drives. NVivo 14 facilitated coding and retrieval.

### Ethics

Approved by Mildmay Uganda Research Ethics Committee (MUREC-2024-451) and Uganda National Council for Science and Technology (SS3374ES). Written informed consent/assent was obtained, with parental consent for minors, in accordance with the Declaration of Helsinki.

### Analysis

Thematic analysis followed Braun and Clarke’s six steps.^19^ Two researchers coded transcripts independently using NVivo 14 (intercoder agreement 85%). Triangulation, member checking, and reflexive journaling enhanced credibility.

### Researcher characteristics and reflexivity

The team included local health professionals and social scientists (two male, two female) experienced in adolescent SRH. Their professional backgrounds and shared language facilitated rapport but could introduce bias. We mitigated this through reflexive journaling, peer debriefing, and independent coding.

### Patient and public involvement

No patient or public involvement in design or conduct; member checking validated findings.

## Results

### Participant demographics

**Table 1.**
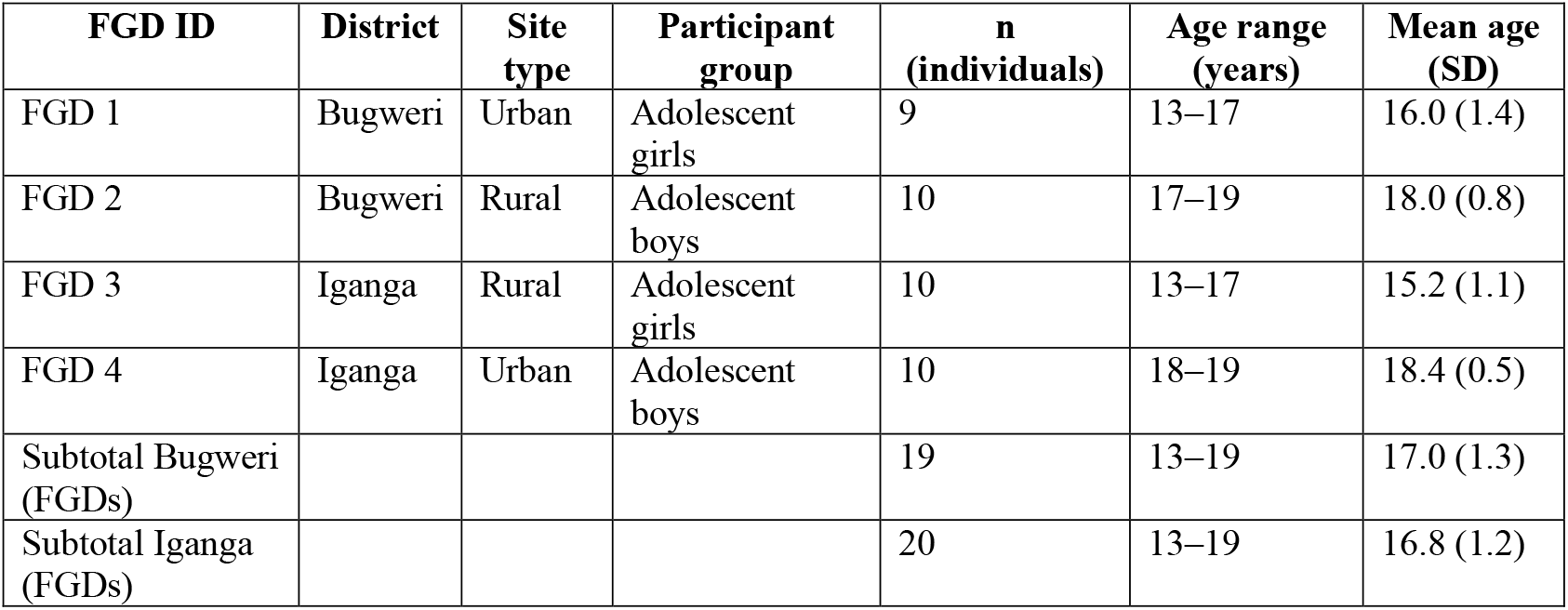
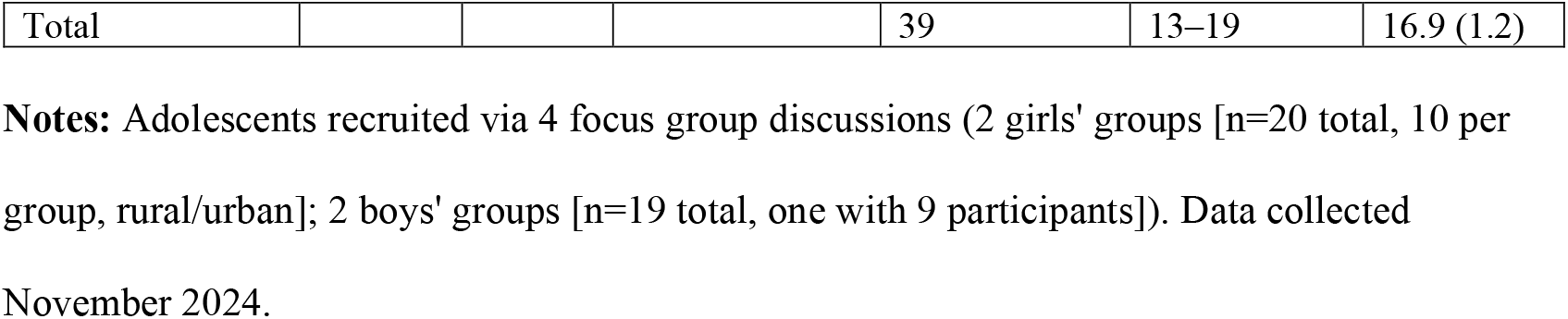
Focus Group Discussions (FGDs) by District and Site.

| FGD ID | District | Site type | Participant group | n (individuals) | Age range (years) | Mean age (SD) |
| --- | --- | --- | --- | --- | --- | --- |
| FGD 1 | Bugweri | Urban | Adolescent girls | 9 | 13–17 | 16.0 (1.4) |
| FGD 2 | Bugweri | Rural | Adolescent boys | 10 | 17–19 | 18.0 (0.8) |
| FGD 3 | Iganga | Rural | Adolescent girls | 10 | 13–17 | 15.2 (1.1) |
| FGD 4 | Iganga | Urban | Adolescent boys | 10 | 18–19 | 18.4 (0.5) |
| Subtotal Bugweri (FGDs) |  |  |  | 19 | 13–19 | 17.0 (1.3) |
| Subtotal Iganga (FGDs) |  |  |  | 20 | 13–19 | 16.8 (1.2) |
| Total |  |  |  | 39 | 13–19 | 16.9 (1.2) |

**Notes:** Adolescents recruited via 4 focus group discussions (2 girls' groups [n=20 total, 10 per group, rural/urban]; 2 boys' groups [n=19 total, one with 9 participants]). Data collected November 2024.

**Table 2.**
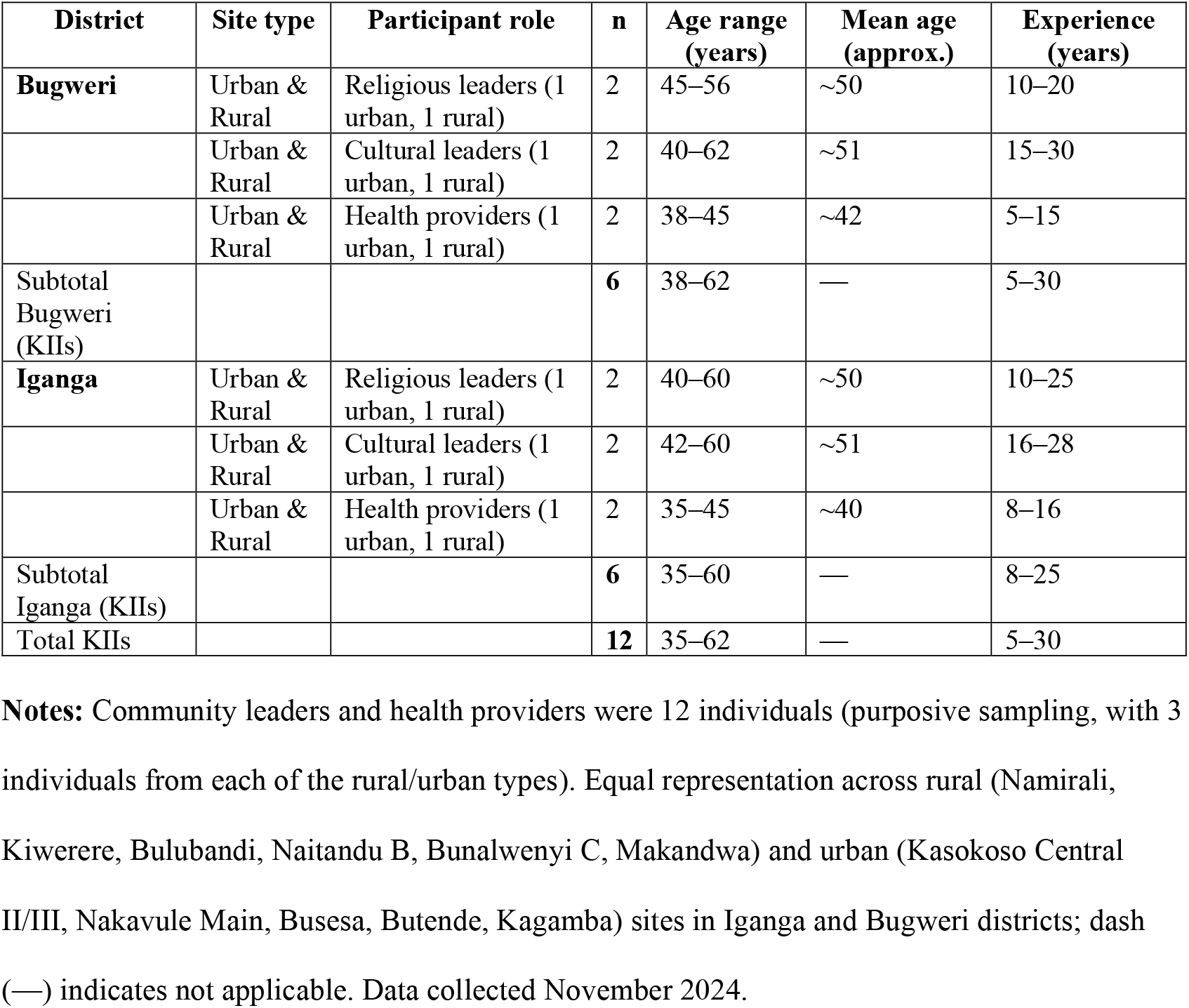
Key Informant Interviews (KIIs) by District and Site.

#### Emergent themes

##### Positive attitudes towards prevention amid self-discovery

Adolescents consistently described SRH services as essential for preventing unintended pregnancies and sexually transmitted infections and for enabling future aspirations, while noting limited parental guidance.

*‘It helps us to prevent sexually transmitted infections and to plan for the future’* (girl, FGD, Bugweri).

Key informants confirmed this preventive motivation: ‘*Most of them, especially girls, fear pregnancies and boys fear illnesses like HIV, so they look for condoms and counselling’* (health provider, KII, Bugweri).

##### Misconceptions and negative beliefs

Despite recognising benefits, adolescents reported widespread myths about contraception, particularly fears of infertility, which discouraged service use.

*‘Sometimes family planning methods may cause you to become barren’* (girl, FGD, Iganga). Providers echoed these concerns: *‘Some have other beliefs that when they go for family planning in future, they will not get children’* (health provider, KII, Iganga).

##### Barriers to service access: distance, privacy and provider attitudes

Structural and interpersonal barriers, including long travel distances, extended waiting times and judgemental provider attitudes, were commonly described.

*‘Government health facilities are far and you can wait many hours before seeing a health worker’* (boy, FGD, Iganga).

A provider highlighted similar challenges: *‘Sometimes there are stockouts, and if adolescents do not find the method they want, they may not come back’* (health provider, KII, Bugweri).

##### Preferences for youth-friendly, community-based services

Adolescents expressed strong preferences for free, confidential services provided in schools or community settings, with peer involvement and gender-matched providers.

*‘During school holidays, services should come to the community and be led by youth because we believe our peers’* (girl, FGD, Iganga).

Health workers endorsed this approach: ‘*We need to collaborate with schools and create adolescent-friendly corners so that young people can pick what they need in privacy’* (health provider, KII, Iganga).

##### Sociocultural influences and system-level factors

Early marriage norms, religious restrictions and inconsistent parental involvement shaped adolescents’ attitudes and access to care, while commodity shortages and limited staff training constrained service delivery.

*‘Parents have lost interest in their children, so the children are growing themselves’* (health provider, KII, Iganga).

A religious leader observed the dual role of faith: *‘Religion can guide good conduct, but sometimes we marry off adolescents early and they miss many opportunities’* (religious leader, KII, Bugweri).

Overall, adolescents demonstrated strong motivation to protect their sexual and reproductive health but faced intersecting sociocultural and system-level barriers. These findings underscore the need for integrated, youth-friendly SRH services that combine accurate information, confidentiality, community engagement and reliable supplies.

## Discussion

This qualitative study explored adolescents’ and key community stakeholders’ perspectives on sexual and reproductive health (SRH) services in Busoga, Uganda. Five interrelated insights emerged.

First, attitudes and beliefs revealed a paradox. Adolescents valued SRH services for preventing pregnancy and sexually transmitted infections, yet persistent misconceptions, particularly fears of infertility from modern contraception, discouraged use. Such myths have been widely documented in Uganda and across sub-Saharan Africa and are known to reduce uptake of family planning methods. ^20 21^

Second, perceptions of services highlighted structural and interpersonal barriers. Cost, distance and negative provider attitudes, including judgemental behaviours and concerns about confidentiality, limited access, especially in rural settings. These barriers reflect findings from Kenya and Ethiopia, where adolescents avoid services when privacy is not assured or when they expect moral scrutiny. ^5 7 22^

Third, service preferences were strikingly consistent. Adolescents expressed a strong desire for free, integrated, peer-led, school or community-based services that offer confidentiality and social engagement. Such preferences expand the World Health Organisation’s adolescent-friendly service standards ^23^ and align with evidence of successful peer education and digital outreach interventions in other low and middle-income settings. ^24 25^

Fourth, sociocultural influences, including early marriage norms, restrictive religious teachings, entrenched myths and inconsistent parental involvement, further constrained SRH service use. While some religious leaders provided positive guidance, others reinforced prohibitions on contraception. Engaging faith leaders and parents is therefore critical to dispel misinformation and support adolescents’ SRH needs. ^26^

Finally, provider and system factors, such as shortages of SRH commodities, limited staff training and weak collaboration across sectors, impeded service delivery. Participants advocated for continuous school-based sensitisation, community outreach and reliable supply chains to improve service uptake.

### Comparison with existing evidence

These findings are consistent with Ugandan research documenting provider stigma and limited adolescent-friendly services. ^20 21^ Regional studies from Kenya and Ethiopia similarly emphasise the importance of confidentiality, affordability and respectful care in adolescent SRH use. ^5 7 22^ The strong preference for peer-led, recreation-integrated approaches builds on WHO adolescent-friendly standards ^23^ and reflects the success of peer education and digital outreach programmes reported in other low and middle-income settings. ^24 25^

Interpreted through the Health Belief Model, adolescents’ high perceived benefits, such as pregnancy prevention, coexist with high perceived barriers such as stigma and cost, which explains their ambivalence towards service use. Andersen’s Behavioural Model likewise highlights how predisposing factors such as gender norms and religious teachings, enabling resources such as transport and free services, and perceived need converge to shape health service utilisation.

### Implications for policy and practice

Addressing sociocultural norms, including early marriage expectations and religious prohibitions, requires sustained dialogue with parents, faith leaders and cultural authorities, supporting calls for multi-stakeholder partnerships. ^26^ Policy actions should prioritise:

- Adolescent-friendly infrastructure and confidential service delivery
- Peer-led outreach integrated with schools and community programmes
- Integration of SRH with other adolescent services, such as mental health support and recreational activities
- Strengthened supply chains and provider training to ensure non-judgmental, youth-centred care

Collectively, these strategies can reduce misconceptions, overcome structural barriers and foster a supportive environment for adolescents to access and use SRH services in Busoga and similar low-resource settings.

## Strengths and limitations

Methodological strengths included triangulation of data from adolescents, health providers, and community leaders; and independent coding with high intercoder agreement, all of which enhanced credibility and transferability. Limitations included the study’s confinement to two districts within the Busoga region, self-report bias, translation from Lusoga to English, and researcher positionality, as local health professionals could have influenced participant responses despite mitigation strategies.

## Future research

Implementation research is needed to test peer-led and school-based SRH interventions and to evaluate digital and community-embedded strategies that could complement facility-based services.

## Conclusion

Adolescents in Busoga navigate SRH amid misconceptions, stigma, and access barriers. Strengthening partnerships, dispelling myths, and allocating resources for adolescent-friendly services are critical to improving utilisation.

## Acknowledgements

We thank all participants and the Iganga and Bugweri District Health Offices.

## Data availability

De-identified transcripts are available from the corresponding author on reasonable request and ethics approval.

